# Development and Validation of the Overall Fidelity Enactment Scale for Complex Interventions (OFES-CI)

**DOI:** 10.1101/2023.02.01.23285328

**Authors:** Liane R Ginsburg, Matthias Hoben, Whitney Berta, Malcolm Doupe, Carole Estabrooks, Peter G. Norton, Colin Reid, Ariane S. Massie, Adrian Wagg

**Affiliations:** School of Health Policy & Management, Faculty of Health, York University, Toronto, Ontario, Canada, M3J 1P3; Faculty of Nursing, University of Alberta, Edmonton, Alberta, Canada, T6G 1C9; Institute of Health Policy, Management & Evaluation, Dalla Lana School of Public Health, University of Toronto, Toronto, Ontario, Canada, M5T 3M6; Rady Faculty of Health Sciences, Max Rady College of Medicine, University of Manitoba, Winnipeg, Manitoba, Canada, R3E 3P5; Cumming School of Medicine, University of Calgary, Calgary, Alberta, Canada, T2N 4N1; School of Health and Exercise Science, University of British Columbia Okanagan, Kelowna, British Columbia, Canada V1V 1V7; School of Kinesiology & Health Science, Faculty of Health, York University, Toronto, Ontario, Canada, M3J 1P3; Division of Geriatric Medicine, Faculty of Medicine & Dentistry, University of Alberta, Edmonton, Alberta, Canada, T5G 2P4

**Author notes:** **Corresponding author:** Liane R Ginsburg.

**Keywords:** Fidelity, trial evaluation, measurement

## Abstract

**Background:** Assessing fidelity in complex interventions is important. Ignoring it increases the risk of discarding potentially effective interventions that failed to work because they were not implemented as intended or, accepting ineffective interventions whose effects were brought about by factors other than the intervention. Unlike simple interventions, fidelity assessment in complex interventions is underexplored and not straightforward. Adapting the evaluative approach used in Objective Structured Clinical Exams (OSCEs), we describe the development and validation of an easy to use, objective approach to fidelity assessment – the Overall Fidelity Enactment Scale for use in complex interventions (the OFES-CI).

**Methods:** We developed the OFES-CI and assessed its accuracy using secondary data collected as part of the process evaluation of the SCOPE intervention. Specifically, we compared the OFES-CI ratings obtained from trained study observers during SCOPE intervention workshops to more detailed and comprehensive qualitative fidelity enactment data collected between and during the same workshops (our gold standard).

**Results:** The OFES-CI demonstrates acceptable reliability against our gold standard assessment approach and addresses the practicality problem in fidelity assessment by virtue of its suitable implementation qualities (acceptability, ease of completion, low burden, low training requirements). It can be easily adapted to assess enactment of a variety of complex interventions.

**Conclusions:** Global ratings approaches validated in the context of assessing clinician competence in OSCEs provide a novel, practical and underexplored opportunity to assess fidelity enactment in complex trials. Given important questions that are increasingly being asked regarding evaluation of complex pragmatic interventions, the proposed approach to fidelity assessment, if incorporated into trial evaluation, stands to provide valuable insights not only into why an intervention may succeed or fail but also how we might carry out adaptations of complex interventions when they are needed.

KEY ARTICLE MESSAGES

- There is a growing knowledge base regarding how to assess the fidelity with which complex interventions (trials) are *delivered*, though there is relatively little knowledge regarding how to efficiently assess the fidelity with which they are implemented (*enacted*) by intervention participants. Data on fidelity enactment is critical for proper interpretation of trial outcomes.
- The present study provides an easy-to-use, valid and reliable approach to assessment of fidelity enactment for use in complex trials (the OFES-CI) and outlines *specific* procedures for assessing fidelity.
- The OFES-CI can be easily adapted for practical assessment of fidelity of other complex interventions. Such fidelity data can help address well known problems with trial replication by providing valuable insight into why interventions succeed or fail and what adaptations may be needed to promote greater success.

## BACKGROUND

When an evaluation shows an intervention did not achieve its aims, its often hard to know if this means the intervention is ineffective or it was simply not implemented as intended. Fidelity of an intervention reflects the extent to which that intervention is implemented as intended^1^ and its assessment is extremely important as it improves our ability to draw conclusions about intervention effectiveness.^2–4^ Ignoring fidelity increases the risk of discarding potentially effective interventions that failed to work because they were not implemented as intended or, accepting ineffective interventions whose effects were brought about by factors other than the intervention.

With some interventions, assessing fidelity is straightforward. For example, in a trial of an order set to improve care for patients with diabetes and related complications, we would assess fidelity simply by looking at how often the order set was used for eligible patients. Assessing fidelity is not so straightforward in trials of complex interventions^3^ where there may be multiple interacting components and multiple actors, such as a trial of team-based PDSA cycles to improve care for nursing home residents. In these trials, assessment of the fidelity with which intervention activities are enacted is itself complex (fidelity enactment^5^ refers specifically to the performance of intervention skills / implementation of core intervention components in the intended situation – see note^1^). Unlike an order set trial which targets individual behaviour and where fidelity is documented on the order set, a more complex trial like a team-based PDSA approach to improve care in nursing homes may or may not produce documented evidence of PDSA implementation that can be used to assess fidelity. Moreover, should such documentation exist, it is unlikely to reflect the nuances of PDSA work or how well (or poorly) the PDSA cycles were done. Given the complexity of more intricate trials, audio or video recording and coding is generally recognized to be the gold standard for assessing fidelity.^6^ Expert assessment of recorded implementation activities is, however, costly, and infeasible with many complex interventions, particularly pragmatic interventions taking place to community settings. Fidelity in complex trials is also sometimes assessed using detailed self-report checklists containing items that reflect core components of the intervention. While practical, self-report approaches are highly susceptible to social desirability bias.

The need for quick and efficient,^7^ psychometrically sound and practical approaches to assessment of fidelity enactment has been highlighted by several recent reviews.^2,3,6,8^ as has the need for studies that outline *specific* procedures for assessing fidelity.^2,9,10^ This study builds on our previous work^11,12^ and describes the development and validation of an easy to use, objective approach to fidelity assessment – the Overall Fidelity Enactment Scale for use in complex interventions (the OFES-CI). Our development and validation work was carried out in the context of the SCOPE intervention (described elsewhere).^13^ SCOPE is a complex intervention conducted in Western Canada in 2019 which aimed to achieve quality improvement in nursing homes using the Breakthrough series model.^14^ SCOPE teaches teams led by healthcare aides to use PDSA approaches to improve resident care (see Box 1 for a description of the SCOPE intervention and schematic). During the one-year SCOPE intervention teams participated in quarterly learning congresses where the PDSA approach was taught (learning congress 1) and reinforced (learning congress 2). Teams were expected to implement PDSA cycles between learning congresses with internal facilitation from local facility leaders, and QI-specific facilitation from an external quality advisor. Teams presented their PDSA implementation progress at learning congresses 2-4.

Given impracticalities of using audio / video recording or observation/audit of teams in their respective nursing homes as they apply discrete steps in the PDSA process, we devised an innovative approach to efficiently assess fidelity to a complex intervention. The approach is an adaptation of the evaluative approach used in Objective Structured Clinical Exams (OSCEs). OSCEs are routinely used to assess competency of health professional trainees prior to entry to practice. In an OSCE, trainees interact with standardized patients in a series of 5–10-minute encounters during which the trainee must assess or resolve a clinical problem. These encounters are observed and evaluated by clinicians who rate the level of competency that the trainee demonstrates during the encounter. The SCOPE intervention requires teams to acquire and apply specific QI skills. Because it is impractical to assess fidelity by observing teams as they apply these skills on a day-to-day basis, we treated their learning congress progress presentation like a standardized patient encounter and applied a similar evaluative approach. We had expert raters observe the progress presentations, ask clarification questions, and then rate fidelity enactment using the OFES-CI tool we developed. Like the OSCE, where competency is typically assessed using global rating scales with explicit criteria, the OFES-CI is a global fidelity assessment scale.

Our approach is supported by the OSCE literature which has shown that (a) subject-matter experts *are* able to reliably evaluate holistic skills in the context of brief, time-limited interactions,^15,16^ and (b) global assessment scales are appropriate when mastery of parts (i.e., discrete skills on a checklist) does not necessarily indicate competency of the whole. Our approach is also supported by the idea that assessing fidelity becomes more difficult as an intervention becomes less prescriptive and expert raters, given their experience, can appropriately use discretion to accept minor variations on intervention fidelity.^17^ We are not the first to use expert raters to assess fidelity. They have been used to assess fidelity *delivery* in psychology and counselling research where interventions tend to involve delivery of complex treatment regimens.

Borrowing from the OSCE literature, drawing on robust psychometric and pragmatic aspects of its approach to competency evaluation, the current paper has two objectives:

### Objective 1

Describe the development of a practical overall measure of fidelity enactment for use in complex trials (the OFES-CI)

### Objective 2

Validate the OFES-CI by comparing its results with fidelity enactment data from a detailed process evaluation as the gold standard

## METHODS

We developed the OFES-CI and then assessed its accuracy using secondary data collected as part of the process evaluation of the SCOPE intervention. Specifically (and described in detail below), we compared the OFES-CI ratings obtained from expert raters who observed SCOPE workshop progress presentations to more detailed and comprehensive qualitative fidelity enactment data collected throughout the 1-year SCOPE intervention (our gold standard).

### Development of the OFES-CI

To achieve objective 1, the OFES-CI was developed following steps outlined by Walton and colleagues^6^ for developing high quality fidelity measures. We also adhered to practices used in our previous work on fidelity assessment^12,18^ and on the use of expert raters.^19^ As a first step, the core components of the SCOPE intervention (Box 1) were analyzed to identify those components that were intended to be enacted by SCOPE intervention participants (unit-based teams led by healthcare aides were the target intervention participants in SCOPE). We proceeded to develop an overall single-item measure of fidelity enactment, the OFES-CI, that reflected the core components of SCOPE and used a 5-point response scale. A rating of “0” indicates ‘*NO / VERY LOW ENACTMENT of SCOPE activities appropriate for [LC#] / inappropriate activities implemented’* and a rating of “4” indicates ‘*VERY HIGH ENACTMENT - extensive implementation of SCOPE activities for [LC#]’*. In keeping with the OSCE assessment approach, the OFES-CI provided ‘Guidelines’ for rating fidelity enactment that included a definition of fidelity enactment, the five-category rating scale, and a few ‘look fors’that reflected the upper two categories on the rating scale (see Figure 1). We obtained feedback about the content and wording of the measure, including the guidelines and ‘look fors’, and pilot tested these (described below). Although this kind of global rating has some degree of subjectivity, OSCE assessment research has shown that global measures are valid and have acceptable reliability, provided the instructions and evaluation criteria are clear and raters are sufficently trained and calibrated.^16^

**Figure 1.**
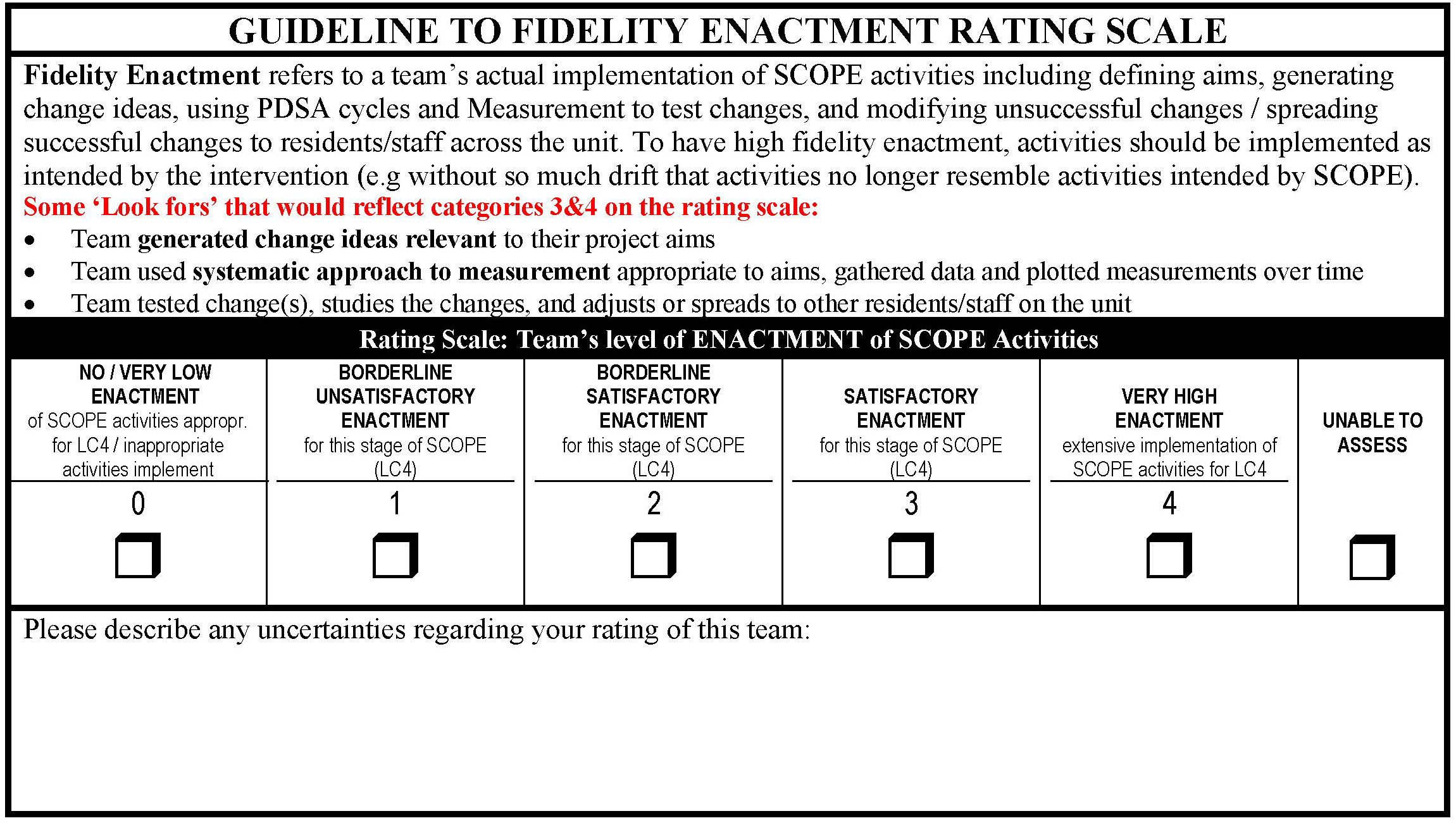
The OFES-CI Global Fidelity Enactment Measure. This fidelity rating scale was applied to project presentations teams gave at learning congresses 2-4. Teams gave a 6-minute presentation in which they described their PDSA activities. Expert raters watched the presentation and had an opportunity to ask the team a question prior to providing a fidelity rating.

### Data Collection Requirements using the OFES-CI

For experts to rate fidelity using the OFES-CI we had to provide opportunities in SCOPE for teams to demonstrate the extent and ways in which they had implemented the core intervention components. As noted above and in the Box 1 schematic, the SCOPE intervention included four, quarterly, learning congresses and at the 2^nd^, 3^rd^, and 4^th^ congresses each team gave a 6-minute ‘progress presentation’ where they were asked to share what improvement activities they had undertaken during the previous quarter, including details of the PDSA work they carried out, and data they collected to know whether their efforts were leading to improvement. We treated these learning congress progress presentations like a standardized patient encounter and applied a similar evaluative approach – expert raters observed the 6-minute progress presentation made by each team and then completed the OFES-CI (figure 1) based on their observations.

Expert raters were members of the SCOPE investigator team from different provinces with expertise in geriatrics, implementation science, and/or improvement science. They were all familiar with SCOPE, QI and the concept of fidelity enactment. Because the learning congresses took place across two large provinces, different experts often attended each congress. As noted, the use of this expert group of raters is supported by OSCE research showing that, when completed by faculty experts, global ratings may have higher reliability and may be more sensitive to variation in intervention participant skills than checklists.^15,20^

### Pilot Testing and Rater Training

We pilot tested the OFES-CI at the second learning congress by having two of our experts provide an enactment rating based on each SCOPE team’s progress presentation. Prior to the congress, all raters participated in a 30-minute zoom training session led by the first author to ensure raters had a common understanding of the ‘Guidelines for rating’ (including the definition of fidelity enactment in SCOPE, the rating scale categories, and the ‘look fors’). Interrater reliability was assessed using a one-way random effects consistency ICC (appropriate when the same raters are not used across all subjects) and was found to be good, bordering on excellent^21^ (ICC=0.73). We therefore proceeded with a single expert rater at the third learning congress.

Following the third congress, during debriefing, experts noted it was sometimes difficult to assess fidelity enactment based solely on the presentation content. Accordingly, we added a short Q&A following each progress presentation at the fourth learning congress and we encouraged raters to ask a question of the team to better enable them to assess fidelity enactment. In the third learning congress debrief we also noticed some raters were overly strict in assessing how well teams carried out measurement in a PDSA cycle. We therefore added a calibration scenario to learning congress four rater training to promote a more widely shared understanding of what would constitute fidelity enactment at the fourth and final learning congress. Finally, so that raters did not feel overly constrained by the 5-point rating scale, they were allowed to provide a .5 rating (between 2 categories). They were also asked to indicate either “I would leave this rating as is”; “I might raise it ½ or 1 category”; or “I might lower it ½ or 1 category”. A comment box was also included where raters could provide commentary to qualify or explain the rating provided.

### Sample

Twenty-seven of the 31 SCOPE teams attended the final learning congress. Global fidelity enactment ratings were collected for each of the 27 teams using the final OFES-CI. Ratings were provided by five experts who attended a final learning congress and who were trained in the manner described above. Each expert provided ratings for 3-7 teams. OFES-CI data were collected at the last learning congress because it marked the end of the one-year trial.

### Validating the OFES-CI – Procedures and Analysis

As noted, audio or video recording and coding is generally recognized to be the gold standard for assessing the fidelity with which an intervention is *delivered*; however, there is no such ‘gold standard’ approach to assessing fidelity *enactment*^6^ since researchers cannot normally record and code the continuous intervention *enactment* processes that occur across multiple sites. Accordingly, we validated the OFES-CI against detailed qualitative data collected as part of a concurrent process evaluation of the SCOPE intervention. For each team participating in SCOPE, qualitative process evaluation data were collected throughout the one-year intervention from various sources: (1) diary entries made by Quality Advisors each time they met with teams between learning congresses to provide QI support – entries included facts and impressions about how implementation was going, (2) responses to open-ended questions provided by SCOPE team members and Quality Advisors on learning congress exit surveys, and (3) observations of team dynamics and intervention engagement conducted by trained members of the research team at each learning congress.

Coding of detailed qualitative process evaluation data is an approach which has been used previously to assess PDSA cycle fidelity^22^ and may be the closest we can get to a gold standard approach to assessment of fidelity enactment. We arrived at a ‘gold standard’ fidelity enactment rating using the first three steps below and then validated the OFES-CI ratings from the final learning congress against the gold standard using the 4^th^ step:

#### Step 1

We conducted a calibration exercise using qualitative process evaluation data for three teams, collected during the three-month period leading up to the third learning congress to see if three authors (LG, WB, MH) could independently code the qualitative data using the OFES-CI categories and arrive at consensus. Comparisons between coders indicated that some scale clarifications were required.

#### Step 2

The same three authors independently coded qualitative data for five teams, this time for the three-month period leading up to the final learning congress. The aim, for coders to achieve ratings that were within 1 point of each other on the 5-point OFES-CI scale, was achieved for 4/5 teams. Coders differing by 1.5 points for the 5^th^ team. Inter-rater reliability was examined using two-way mixed consistency average measures Intra-class correlations (ICCs), appropriate for estimating the reliability of the mean ratings provided by the same set of coders for ordinal data.^23^ The ICC was excellent for these 5 sites (0.95), enabling us to proceed to step 3.^21^

#### Step 3

The remaining 22 teams were coded by two of the authors – LG and either WB or MH. Both coders independently applied the OFES-CI to the qualitative data for the 3-month period leading up to the final learning congress then discussed any cases where ratings were more than one point apart. Inter-rater reliability for all 27 teams that participated in the final learning congress (5 teams coded in step 2 and 22 teams coded in step 3) was examined using a one-way random effects average measures ICC, appropriate where teams were not coded by the same coders.^23^ For each team, the two coders’ scores were averaged to create a ‘gold standard’ enactment rating (i.e., an enactment rating based on review of detailed qualitative data on SCOPE implementation activities that took place during the final three months of the intervention).

#### Step 4

The OFES-CI was validated by comparing its results (i.e., from experts at the final learning congress) with the gold standard enactment rating for all 27 teams that participated in the final SCOPE learning congress using a one-way random effects single measures ICC.^24^ These ICC parameters are appropriate given we don’t have a fully crossed design (i.e., not all pairs of ratings were provided by the same coders) and ultimately allows us to quantify reliability based on ratings provided using a single assessment approach (i.e., use of the OFES-CI by a single subject-matter expert in the context of time-limited interaction at the end of an intervention).

For interpretation of all ICCs we used the classification proposed by Cicchetti (inter-rater reliability less than 0.40 is poor; 0.40 - 0.59 is fair; 0.60 - 0.74 is good; 0.75 – 1.00 is excellent).^34^ Walton et al.,^6^ use a threshold of >0.60 as acceptable against a gold standard approach.

## RESULTS

### Generating the Gold Standard fidelity enactment rating

Inter-rater reliability (Step 3 above) was excellent (one-way random effects average measures ICC = 0.93), indicating that coders had high agreement in their application of the OFES-CI to the qualitative data. As a result of this high level of agreement we used the average score provided by two coders as the gold standard fidelity enactment rating for each team.

### Validating the OFES-CI against the Gold Standard fidelity enactment rating

Using all 27 cases (step 4 above), inter-rater reliability performed to assess the degree to which the OFES-CI rating from final learning congress was consistent with the gold standard enactment ratings was ‘fair’ (one-way random effects single measures ICC = 0.58). There was one site with a gold standard enactment rating of 0.25 (“*NO / VERY LOW ENACTMENT of SCOPE activities*”) and an OFES-CI expert rating from the learning congress of 4.0 (“*VERY HIGH ENACTMENT*”). We examined comments from the OFES-CI rating form for this team which stated that “*They are doing gigantic amounts of stuff…but they seem to have done so before SCOPE … I REALLY wonder to what extent we can attribute the good ratings above [the OFES-CI ratings] to SCOPE… [several initiatives described] …were already successful - how much has SCOPE added????*”. Unfortunately, these comments were not reviewed immediately following the learning congress. Had they been reviewed, we would have asked the reviewer to revise their rating to reflect activities *enacted as part of SCOPE*. After removing data from this site, inter-rater reliability was in the ‘good’ range (ICC = 0.71), exceeding the threshold of 0.60 which Walton et al.,^6^ suggest is acceptable against a gold standard approach.

Nine of the 27 OFES-CI ratings included certainty adjustments (recall they could indicate they might raise or lower their rating by 0.5 or 1 category). We examined their effects by adjusting the OFES-CI learning congress rating up or down by half a point for these nine cases. The ICC worsened slightly when these adjustments were included.

As a final analysis, we looked for evidence of any systematic differences between the OFES-CI rating and the Gold Standard fidelity enactment rating (i.e., was gold standard always higher or lower?) and between the five raters. Figure 2 shows the distribution of gold standard fidelity enactment rating and OFES-CI rating *difference scores* for all 26 cases (far left boxplot) and for each rater (excluding the case described above where the OFES-CI measure was incorrect). For all 26 cases, the mean difference between the two ratings is -0.37 (median difference = -0.17) indicating the gold standard ratings (mean=2.25) were, on average, 0.37 points lower than the OFES-CI ratings (mean=2.62). The left boxplot also shows that 75% of the gold standard and OFES-CI ratings were within one point of each other. None of the individual raters OFES-CI ratings were systematically higher or lower than the gold standard rating, although 1/3 of rater 1 and rater 4’s ratings were > 1 point away from the gold standard.

**Figure 2.**
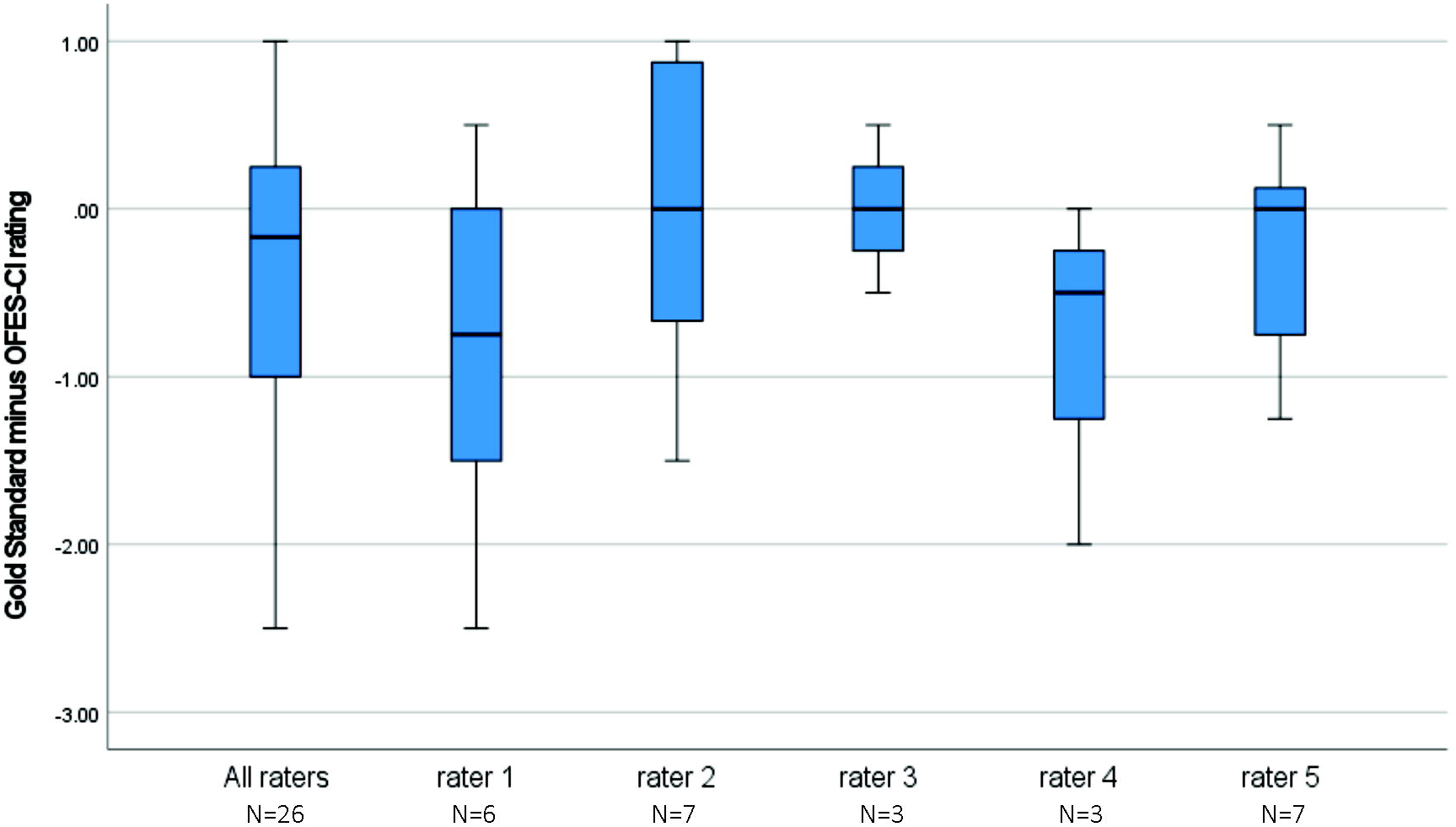
Gold standard fidelity enactment rating and OFES-CI rating Difference Scores.

## DISCUSSION

The need for psychometrically sound and practical/implementable approaches to assessment of fidelity *enactment* has been highlighted by several recent reviews^2,3,6,8^ as has the need for studies that outline *specific* procedures for assessing fidelity.^2,9,10^ This study builds on robust approaches to assessment used in medical education^16^ and describes the development and validation of an Overall Fidelity Enactment Scale for use in complex interventions (the OFES-CI). The OFES-CI demonstrates acceptable reliability against our gold standard assessment approach and addresses the practicality problem in fidelity assessment^18^ by virtue of its suitable implementation qualities (acceptability, ease of completion, low burden, low training requirements). It can be easily adapted to assess enactment of a variety of complex interventions.

Availability of validated tools and practical guidance for their use were identified as key facilitators for enhancing and addressing intervention fidelity in trials by 70-80% of researchers surveyed in a recent study.^25^ The OFES-CI development process we describe is generalizable – it provides a comprehensive approach to the assessment of fidelity enactment in complex trials, including how to adapt the measure to a particular intervention context, and how to pilot test and train expert raters to use it. The OFES-CI approach can meet the needs of researchers by overcoming three practical and methodological challenges associated with assessment of fidelity enactment that have been identified in the literature: (1) the fact that there is no gold standard approach for measuring fidelity *receipt* or *enactment*^6^ (though we contend that collecting and coding detailed process evaluation data may offer one such approach); (2) fidelity enactment has typically been assessed using participant self-report checklists which have been shown to have unclear reliability and validity and low concordance with observer ratings;^8^ (3) fidelity measures, including enactment measures, need to be specific to intervention skills and their measurement properties are therefore rarely established.^6^

Similar to Walton’s (2020) findings regarding the importance of piloting fidelity measures, our piloting, training and calibration work support the importance of these processes in the development and application of the OFES-CI to any complex intervention. Our results showed that rater 1 deviated most from gold standard enactment ratings. This rater was unable to attend the training and although he received a solo makeup training session, it did not allow him to discuss the training scenarios with other raters. Piloting and training may be particularly important for global fidelity enactment measures that assess the enactment of multiple intervention components in a single measure. We concur with Walton’s suggestion that clear definitions must be provided to expert raters in the form of guidelines to make rating easier and limit individual judgement and subjectivity”.^6 p55^

Our validation analysis comparing the OFES-CI ratings to the gold standard (objective 2) identified one large discrepancy, described above, where the OFES-CI rating indicated *very high enactment* while the gold standard rating suggested *no or very low enactment*. Fortunately, the OFES-CI rating form asks raters to describe any uncertainties regarding the rating they provided, which enabled us to resolve the discrepancy. Researchers using the OFES-CI are strongly encouraged to retain the open-text field to permit raters to qualify their ratings if necessary. More importantly, OFES-CI rating forms should be checked by a member of the research team immediately following completion to identify any instances where qualitative comments do not match the rating provided, so that discrepancies can be resolved. We did not do this which resulted in a missing OFES-CI rating for one of the teams in our analysis. Studies of complex group-or organization-level interventions, even large ones, often do not have large samples^26^ and its therefore crucial to minimize missing data.^18^

The OFES-CI requires researchers to build opportunities to assess fidelity enactment into an intervention the way we did in SCOPE – an approach which offers important practical benefits for assessing fidelity enactment, and for trial evaluation more broadly. Using the methods described here for development, data collection, pilot testing, and training, research teams can adapt the OFES-CI to their intervention content and context and can practically evaluate overall fidelity enactment in complex trials. Given robust evidence from the OSCE literature showing that global rating scales can provide a faithful reflection of competency when completed by subject-matter experts in the context of brief time-limited interactions,^15,16^ and the findings of the present study, we are confident that the OFES-CI offers a sound and judicious approach to assessing fidelity enactment that is not currently found in the literature. While continued validation of the OFES-CI approach in other intervention contexts is encouraged, research teams can use the OFES-CI approach to understand and quantify fidelity enactment in complex trials without undertaking the validation procedures and analysis we conducted using the gold standard. Indeed, as noted above, previous work by this team using the OFES-CI approach, without the validation work described here, showed evidence of its predictive validity.^12^ Ultimately, quantitative OFES-CI ratings, together with open-ended comments provided to enrich understanding of enactment, can offer considerable advantages for the analysis and interpretation of trial effectiveness data. To evaluate trials in an even more comprehensive way, it may be most valuable to use the OFES-CI as part of a larger process evaluation.^4^

### Limitations & Future Research

Studies of interrater reliability should be designed so that ratings are independent to avoid inflating ICCs.^27^ In the current validation study, the three authors who coded the qualitative data also provided some of the OFES-CI ratings at the final learning congress. To mitigate potential bias, coders were blind to the team names when coding the qualitative data. Recall bias is further mitigated by the fact that more than two years elapsed between Spring 2019 when OFES-CI ratings were provided at the final SCOPE learning congress and Fall 2021 when qualitative data were coded to obtain gold standard enactment ratings.

The present study focuses on a pragmatic psychometrically sound approach to assessing overall fidelity enactment in complex trials. It does not explore the factors that influence fidelity enactment or their mechanism of impact – explorations which could support fidelity enhancement. Fidelity enhancement could be the subject of future research, perhaps by exploring differences between high and low fidelity enactment sites.

## CONCLUSIONS

Global ratings approaches validated in the context of assessing clinician competence in OSCEs provide a novel, practical and underexplored opportunity to assess fidelity enactment in complex trials. Given important questions that are increasingly being asked regarding evaluation of complex pragmatic interventions,^28^ the proposed approach to fidelity assessment, if incorporated into trial evaluation, stands to provide valuable insights not only into why an intervention may succeed or fail but also how we might carry out adaptations of complex interventions when they are needed.

## Data Availability

All data produced in the present study are available upon reasonable request to the authors provided they adhere to our data confidentiality and access policies.

## DECLARATIONS

### STATEMENT OF ETHICS APPROVAL

This study was approved by the Research Ethics Boards of the University of Alberta (Pro00000012517), University of British Columbia (H14-03286), Operational approval was obtained from all included facilities as required. SCOPE sponsors and team members were asked for informed consent prior to taking part in the study.

### CONTRIBUTORSHIP STATEMENT

…LG is responsible for the overall content as guarantor.

### DATA AVAILABILITY STATEMENT

#### Data are available upon reasonable request

De-identified data specific to this study can be requested through the TREC Data Management Committee on the condition that researchers meet and comply with the TREC and HRDR data confidentiality policies. Data are part of the TREC program of research which has established comprehensive data and intellectual property policies. TREC data are housed in the secure and confidential Health Research Data Repository (HRDR) in the Faculty of Nursing at the University of Alberta (https://www.ualberta.ca/nursing/research/supports-and-services/hrdr), in accordance with the health privacy legislation of participating TREC jurisdictions. The OFES-CI measurement instrument is included in the article.

---

**Box 1. The SCOPE Intervention with Schematic**

- SCOPE is a multicomponent pragmatic trial at the level of the resident care team in 31 nursing homes. SCOPE teaches local HCA-led teams to implement improvement initiatives based on current best evidence. [25] [26] SCOPE is unique in engaging and equipping HCAs to *lead* an improvement team.
- SCOPE is modelled on the Institute for Healthcare Improvement’s Breakthrough Series Collaborative Model[27] and was designed to be *implementable*. Using the PARiHS framework,[28,29] SCOPE addresses technical aspects of conducting a PDSA cycle, provides facilitation, and addresses contextual factors necessary to support implementation.
- SCOPE trial outcomes included best practice use and improvement in the clinical area that teams chose to work on: pain, responsive behaviours, or mobility. Outcomes were measured using Resident Assessment Instrument-Minimum Data Set (RAI-MDS 2.0) indicators. [30]
- The year-long intervention began in June 2018 in four health regions in the Canadian provinces of Alberta and British Columbia). Each of the 31 nursing homes had one unit-based improvement team. Teams had five to seven members, were led by an HCA, and included at least two HCAs.
- Teams attended quarterly learning congresses with other teams in their region to network and participate in short plenary sessions and activities on the improvement model, measurement in PDSA cycles, and team dynamics and function. **Teams presented on project progress at the 2**^**nd**^, **3**^**rd**^ **and 4**^**th**^ **learning congresses**.
- Teams received active support from a *Team Sponsor* (unit-level care manager/clinical educator) and a *Senior Sponsor* (nursing home-level administrator). Teams received coaching from a *Quality Advisor* **(QA)** to support **QI** activities and instil a new approach to improvement work at the bedside. Researchers in geriatrics, nursing, implementation science, **QI**, and health services supported the quality team.
- The core components of the intervention include:
  1. Care aide-led teams working on a focused clinical area
  2. Use of quality improvement methods by unit teams (change concepts, measurement, Plan-Do-Study-Act cycles)
  3. In-person meetings with all teams (quarterly learning congresses (LCs))
  4. Ongoing support from a Quality Advisor during action periods between LCs
  5. Supporting leaders to facilitate and support change and the care aide-led teams

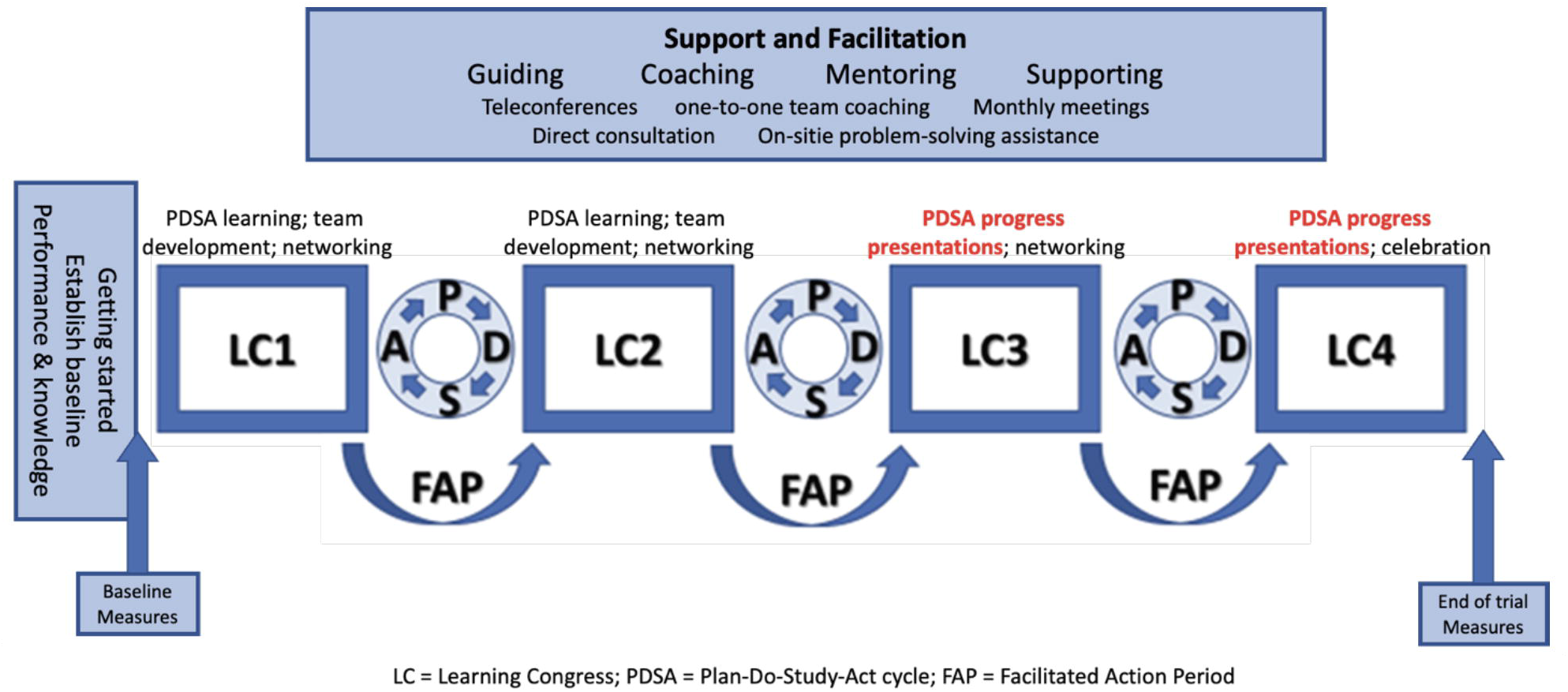

## Footnotes

Note1: We focus here on fidelity *enactment*. Fidelity enactment is often different from fidelity delivery in complex trials and it is crucial given intervention targets must ultimately implement (i.e., enact) the core components of an intervention to achieve complete fidelity. Fidelity enactment is also severely underexplored (Bellg et al., 2004; Hasson, 2010; Toomey et al., 2020; Walton et al., 2017). For a detailed discussion of different types of fidelity (e.g., delivery of an intervention versus enactment of an intervention), different fidelity models, and the fidelity-adaptation debate see Ginsburg et al., (2021).

